# Socio-economic disparities in social distancing during the COVID-19 pandemic in the United States

**DOI:** 10.1101/2020.11.07.20201335

**Authors:** Romain Garnier, Jan R. Benetka, John Kraemer, Shweta Bansal

## Abstract

**Importance:** Eliminating disparities in the burden of COVID-19 requires equitable access to control measures across socio-economic groups. Limited research on socio-economic differences in mobility hampers our ability to understand whether inequalities in social distancing are occurring during the SARS-CoV-2 pandemic.

**Objective:** To assess how mobility patterns have varied across the United States during the COVID-19 pandemic, and identify associations with socio-economic factors of populations.

**Design, Setting, and Participants:** We used anonymized mobility data from tens of millions of devices to measure the speed and depth of social distancing at the county level between February and May 2020. Using linear mixed models, we assessed the associations between social distancing and socio-economic variables, including the proportion of people below the poverty level, the proportion of Black people, the proportion of essential workers, and the population density.

**Main outcomes and Results:** We find that the speed, depth, and duration of social distancing in the United States is heterogeneous. We particularly show that social distancing is slower and less intense in counties with higher proportions of people below the poverty level and essential workers; and in contrast, that social distancing is intense in counties with higher population densities and larger Black populations.

**Conclusions and relevance:** Socio-economic inequalities appear to be associated with the levels of adoption of social distancing, potentially resulting in wide-ranging differences in the impact of COVID-19 in communities across the United States. This is likely to amplify existing health disparities, and needs to be addressed to ensure the success of ongoing pandemic mitigation efforts.

## Introduction

Treatment options and vaccines are under development to address the COVID-19 pandemic^1^. However, when the ability to detect new infections remains limited by testing capacity and the lack of nationwide syndromic surveillance capabilities^2^, non-pharmaceutical interventions represent public health agencies’ only immediate tools to limit outbreak size and spatial scale^3^. In the United States, state and local governments are primarily responsible for measures such as school or business closures^4^. Historically, similar measures have been used to respond to pandemics including during plague outbreaks in the Middle Ages^5^, or during the 1918 Spanish flu pandemic^6^. Data collected during the early part of the ongoing COVID-19 pandemic, in particular on the dynamics of the outbreak in China, indicates that these measures can be successful at limiting COVID-19 outbreak sizes^7^ and at delaying large scale spread^3^.

However, social distancing may occur differently across communities, especially in the United States where sectors including transportation or food retail have lower wages and represent a larger fraction of workers deemed essential than other sectors of the workforce^8^. Assessing this differential impact requires the use of fine-scale mobility data, a stream of information that has proven useful in the early assessment of social distancing measures in the United States^9^, Italy^10^, and France^11^. Digital technologies have taken the center stage in the response to COVID-19^12^, and mobility data in particular has allowed to assess the response to non-pharmaceutical interventions^13,14^.Previous studies report large scale reductions in movements, with numbers quickly reaching values typically observed during holiday periods^9^. Furthermore, in the United States, changes in mobility were associated with reductions in COVID-19 cases^15,16^ and in the reproduction number^17^ and mobility data also revealed that the changes were largely already underway when state or county stay-at-home orders were issued^15,18,19^. Previous studies using mobility data have also suggested the potential for inequalities in the ability to practice social distancing based on income^20^, race and education^19^, or on the availability of healthcare providers^21^. However, most of these studies consider these determinants in isolation and do not allow to disentangle the potential additive effects the socio-economic make-up of counties on the ability of their population to practice social distancing. The focus of these studies has also largely been on understanding how social distancing was influenced by local or regional decisions, and whether socio-economic factors changed the responses to state and local interventions.

Here, we focus on an ecological understanding of how mobility varies with socio-economic characteristics, rather than assessing what drives these changes. As outlined above, multiple studies have aimed at understanding causes of mobility behavior changes. We, however, seek to understand how the patterns of mobility vary across socio-economic characteristics (regardless of the cause) during different stages of the pandemic response. In particular, we ask how quickly, how deeply, and for how long mobility change occurred in locations according to their socio-economic characteristics. Our approach does not seek to differentiate between spontaneous changes in mobility, for instance in response to news coverage, and changes in response to state or county mandated orders. Rather, we focus on the resulting changes in mobility and how these differ by location and socio-economic status.

## Material and methods

To measure mobility, we obtained daily county-specific mobility data for the United States from 24 February to 14 May 2020 through a partnership with Unacast^22^. The dataset is based on the GPS location data collected from applications installed on tens of millions of devices, and complies with the General Data Protection Regulation (GDPR) and the California Consumer Privacy Act (CCPA)^23^. The dataset has been shown to be representative by geographical locations, income levels, sex, and age in an analysis conducted by Unacast^24^. The fraction of all devices observed varies by location and time, and has been captured in our analysis (details below).

As outcome measures, we considered data on the changes of three measures of mobility provided by Unacast: daily distance traveled (hereafter ‘distance traveled’), rate of visitation to non-essential places (hereafter ‘visitation rate’), and rate of encounters between devices within a 50 meters radius within an hour (hereafter ‘encounters rate’). Distance traveled reflects the average distance between the home locations of users and locations visited in a day. Visitation rate reflects the number of visits to non-essential locations; the definition of non-essential venues is based on state-specific guidelines and policies, and includes all locations besides those deemed essential (e.g. food stores, pet stores, and pharmacies; more information can be found on Unacast’s website^25^). Encounters rate measures the likelihood of proximity between any two users within 50 meters over a one hour period.Each change is calculated relative to a county-specific baseline calculated from values obtained during a period of several weeks prior to the onset of major COVID-related changes in mobility in the United States (10 February 2020 to 4 March 2020 for distance traveled and and visitation rate; 24 February 2020 to 4 March 2020 for encounters rate). The resulting dataset covers 3054 counties for distance traveled and encounters rate and 2067 counties for the visitation rate. The county-level data on each measure described above can be accessed by contacting Unacast^22^, and our model-processed data and the code for the analysis are available on Github^26^.

In order to summarize the mobility time series, using fbprophet^27^, we first fit a non-linear model to the county-level social distancing time series, including a weekly trend to account for workweek variation. In essence, the fbprophet algorithm uses a Generalized Additive Model (GAM) approach to fit a piecewise regression. It allows setting the number of potential changepoints, and makes it possible to assess where breaks in the trend occur over the course of the social distancing time series. In most counties the underlying trend can be separated into four phases (i.e. four breaks in the trend): (i) baseline period; (ii) period of entry into social distancing, measured by the rate of mobility decrease; (iii) social distancing period, described by a sustained reduction in mobility outcomes; (iv) period of exit from social distancing, measured by the rate of mobility increase after sustained social distancing. This dynamic is specific to 2020; these four phases are not evident in 2019 (Figure S1). From these model fits, we extracted several values that allow to characterize the changes in mobility during entry into and exit from social distancing, as well as the mobility level during the period of sustained mobility reduction. Counties with no detectable increase in mobility after sustained social distancing (i.e. where the slope of the trend at the end of the time series remains non-positive) were not included in the phase iv analysis. Each county time series is thus summarized by four values (or three values for the counties with no detectable phase iv increase), which are used in the statistical analysis. We do not distinguish whether these changes are spontaneous in response to COVID-19 or occur in response to public health policies; we only define the phases based on changes in mobility rates. That is, we do not seek to explain why mobility changed but rather how it changed.

Our exposures of interest relate to the socio-economic composition of each area: racial composition, population density, proportion living below the poverty level, and proportion of the workforce in industries designated as essential. Thus our exposures of interest are area-level features, not individual features. We obtained information on racial composition and population density from the 2018 American Community Survey^28^, and on the proportion of people below the poverty level from the Small Area Income and Poverty Estimate program^29^. We estimated the proportion of workers in industries designated as essential^30^ from the Quarterly Census on Employment and Wages for the fourth quarter of 2019^31^.

We also adjust for the fraction of devices observed in each county because sampling of mobile devices tends to vary geographically and over time.

We ran linear mixed models to analyze the associations between the social distancing summary values and the socio-economic variables, with state as a random effect and using the standard 0.05 significance threshold. In the main analysis, we investigated independent associations with each covariate. We added a post-hoc secondary analysis in which interaction terms between covariates were added to elucidate findings from the main analysis. As a sensitivity analysis, we fit an alternative model with state as a fixed rather than a random effect in order to adjust for any additional unmeasured state-level features. All analyses were run in Python 3.6.

## Results

Social distancing is heterogeneous at the county level (Figure 1). In counties with a higher proportion of people in poverty, social distancing was weaker: mobility was less restricted during the period of sustained mobility reduction and the change was slower to take place during the period of entry into social distancing for all three measures of mobility; additionally, the resurgence in mobility was faster during the period of exit from sustained reductions in mobility for two of three measures of mobility (Figure 2). Counties with a higher proportion of essential workers saw weaker social distancing for all mobility markers during the period of sustained mobility reduction and a slower entry into social distancing based on two mobility markers. The rate of exit from social distancing is less predicted by the proportion of essential workers.

**Figure 1:**
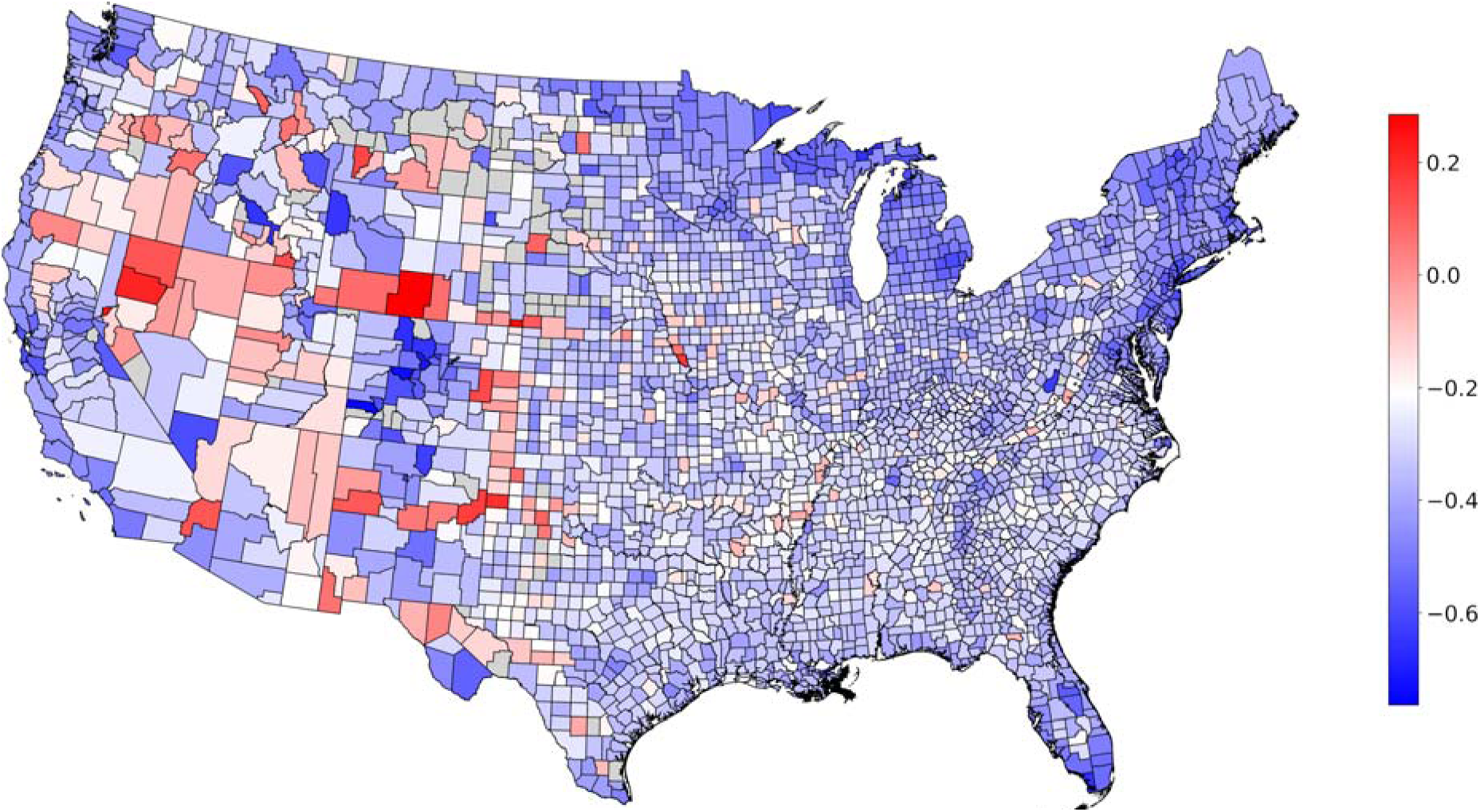
Heterogeneity in mobility during social distancing. Map shows average mobility during COVID-19 social distancing at the county level in the (continental) United States, relative to the pre-COVID-19 baseline. A positive value indicates an increase in distance traveled, and a negative value a decrease in distance traveled. Counties for which the data is not available are shown in grey. We note that the colormap is centered at the 90% percentile of the decrease in mobility.

**Figure 2:**
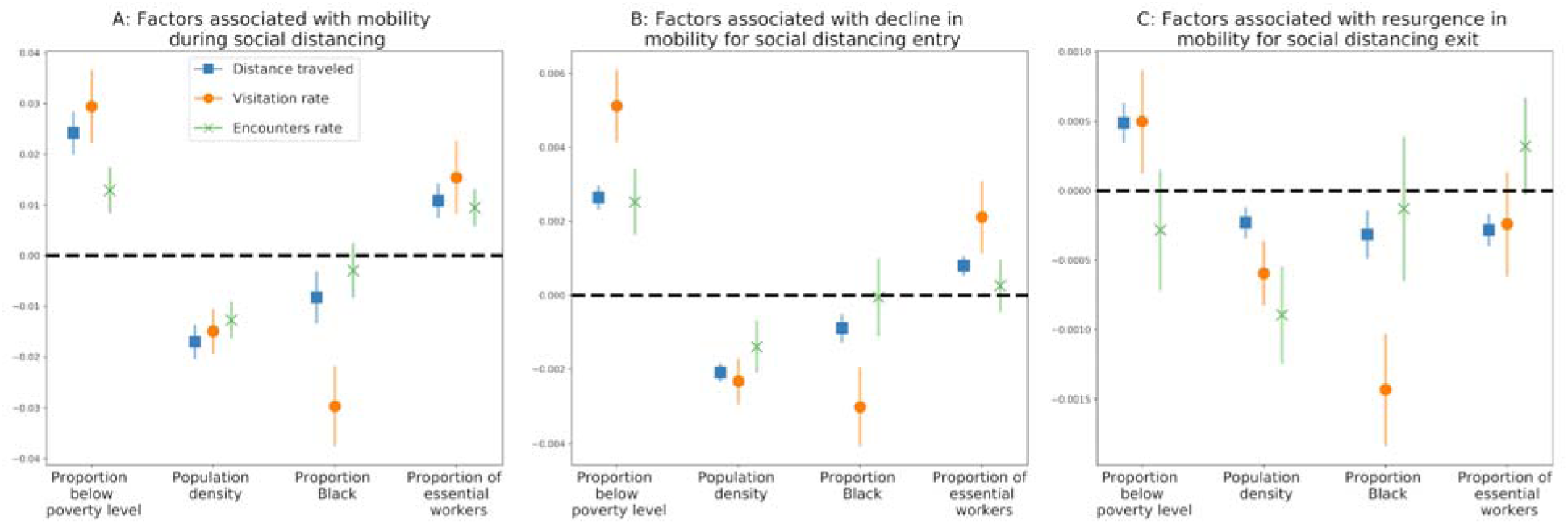
Socio-economic factors associated with social distancing. We show the regression coefficients for four socio-economic predictors (A) mobility during social distancing (phase iii); (B) decline in mobility during entry to social distancing (phase ii); and (C) resurgence in mobility during exit from social distancing (phase iv). Results are shown for three measures of mobility: distance traveled, visitation rate, and encounters rate. The marker denotes the mean coefficient, and error bars show the 95% confidence interval. A positive association (above the dashed line) indicates that an increase in a given factor leads to a weaker implementation of social distancing. A negative association (below the dashed line) indicates that an increase in the given factor is associated with a stronger implementation of social distancing.

Contrastingly, in counties with a larger proportion of Black individuals or a higher population density, social distancing is stronger, with a faster entry into social distancing, lower mobility levels during the period of sustained mobility reduction, and a slower resurgence during the period of exit from social distancing (Figure 2). Results are not significant for the encounter rate for the proportion of Black population. Full statistical details are provided in Table S1.

When interaction effects are added to the model of distance traveled in the period of entry into mobility reductions, all variables follow the same qualitative and quantitative patterns (Table S2). We find a significant positive interaction between the proportion of essential workers and Black population, and a negative interaction between essential workers and low-income workers.The interaction between the proportion of Black people and of low-income workers is not significantly associated with mobility.

Substituting the state random effect for a fixed effect yielded very similar results. There were only three differences of note: the association between the mobility resurgence measure as the visitation rate in the period of exit from social distancing and the proportion of low income workers became non-significant, while the associations between the resurgence in encounters rate in the period of exit from social distancing and the proportions of essential and of low-wage workers became significant.

## Discussion

The COVID-19 pandemic has highlighted significant health disparities in the United States, similar to the pre-existing health inequities driven by income inequality and racial injustice that existed in the country^32^. Understanding the role of behavioral interventions in driving variation in COVID-19 burden is crucial to our current and future outbreak response. Our study shows that changes in interaction in response to the pandemic is geographically heterogeneous and associated with county-level socio-economic factors. This is true for both the level of mobility restriction implemented during the social distancing phase, but also for the rate at which populations enter (“response engagement”) and exit (“response fatigue”) the social distancing phase.

Our analysis reveals that the occupational make-up of counties is associated with how deeply and for how long social distancing is maintained. Populations consisting of more essential workers, who have maintained food service, public transportation and healthcare services during the pandemic^8^, understandably participate less in social distancing, experiencing greater risk. Additionally, lower-income populations participate less in social distancing, likely in part because low-wage workers may have less access to job protections or paid leave. Our results may help explain why lower-income counties have suffered a disproportionately high death burden from COVID-19^16^, but also need to be taken in light of the more general role that low income plays on negative health outcomes^20^. Reduced access to employer-sponsored healthcare^33^ could further limit testing and treatment-seeking behavior, and potentially worsen outbreaks in these communities. In rural (low population density in our study) communities, the need to travel farther to access essentials such as food or healthcare^34^ may also limit social distancing, and would further confirm the existing disparity whereby rural counties suffer from poorer health outcomes than their more urban counterparts^35^.

Importantly, we also find that counties with larger Black populations show stronger social distancing during all phases, after controlling for the effect of income, occupation, and density. Our finding is supported by more local observations of differences between predominantly Black and White neighborhoods, for instance in Detroit^36^. We also find that social distancing remains more limited where high proportions of Black population are combined with high proportions of essential workers, possibly because minorities may be overrepresented in certain essential occupations^8^. Despite stronger distancing, there is growing evidence that African-American communities experience higher rates of infection and deaths from COVID-19^16,37,38^. We advocate for additional work on the structural racism that is at the root of such health disparities^39^ and on the role of privilege in the differential burdens imposed by COVID-19 on a variety of communities^21,40^.

There is a risk of the ecological fallacy if our results are interpreted as applying to individuals with the attributes we investigated rather than the share of attributes in communities. Survey and qualitative studies would help explain how individual, community, and public policy-level factors explain these associations.

Without large-scale test-trace-isolate programs or other interventions, intermittent social distancing will continue to be needed to contain cases and minimize the strain on health systems^41^. Technological solutions are being suggested and to an extent implemented^12–14^, but are not without their limitations. The large-scale use of mobility data and other digital technologies (e.g. for contact tracing) has opened up a debate on the responsible use of these emerging data stream^12,42^, in order for instance to ensure that privacy concerns are properly assessed and taken care of. These technologies would also likely be most effective with a spatially and socially homogeneous testing strategy. Similarly, the long-term success and equity of a mitigation strategy hinges on paying more attention to the geographic heterogeneity in outbreak mitigation, and a focus on the role of social and employment policies that affect the ability of individuals to engage in behavioral interventions.

## Data Availability

The county-level time-series data on each measure of social distancing used in the manuscript can be accessed by contacting Unacast, and our model-processed data and the code for the analysis are available on Github.

https://github.com/bansallab/SESdistancing

## Acknowledgements

Research reported in this publication was supported by the National Institute Of General Medical Sciences of the National Institutes of Health under Award Number R01GM123007. The content is solely the responsibility of the authors and does not necessarily represent the official views of the National Institutes of Health. The authors declare no conflict of interest.

The data and the code used for the statistical analysis are available at https://github.com/bansallab/SESdistancing.

