## Supplementary table and figures for "Socio-economic disparities in social distancing during the COVID-19 pandemic in the United States"

Romain Garnier, Jan R. Benetka, John Kraemer, Shweta Bansal

**Supplementary material**

Table S1: Statistical details for the linear mixed models. For each combination of predictor, response variable, and social distancing measure, we provide the t-value and the p-value.

| Model | Variable | Distance traveled | | Visitation rate | | Encounters rate | |
|---|---|---|---|---|---|---|---|
| | | ***t*** | ***p*** | ***t*** | ***p*** | ***t*** | ***p*** |
| **Mean value in phase iii** | Proportion poor | 11.1 | <0.001 | 7.92 | <0.001 | 5.51 | <0.001 |
| | Proportion essential | 6.19 | <0.001 | 4.15 | <0.001 | 5 | <0.001 |
| | Population density | -9.85 | <0.001 | -6.52 | <0.001 | -6.8 | <0.001 |
| | Proportion Black | -3.15 | 0.002 | -7.39 | <0.001 | -1.08 | 0.28 |
| **Slope in phase ii** | Proportion poor | 16.1 | <0.001 | 10.2 | <0.001 | 5.6 | <0.001 |
| | Proportion essential | 6.05 | <0.001 | 4.21 | <0.001 | 0.72 | 0.47 |
| | Population density | -16 | <0.001 | -7.52 | <0.001 | -3.88 | <0.001 |
| | Proportion Black | -4.48 | <0.001 | -5.58 | <0.001 | -0.1 | 0.92 |
| **Slope in phase iv** | Proportion poor | 6.65 | <0.001 | 2.6 | 0.009 | -1.28 | 0.2 |
| | Proportion essential | -4.78 | <0.001 | -1.26 | 0.21 | 1.79 | 0.07 |
| | Population density | -3.98 | <0.001 | -5.05 | <0.001 | -4.99 | <0.001 |
| | Proportion Black | -3.61 | <0.001 | -6.95 | <0.001 | -0.49 | 0.62 |

Table S2: Statistical details for a linear mixed model of distance traveled and the mean value in phase iii including interactions between the proportion of Black population and the poverty status, the proportion of Black population and the essential status, and the poverty status and the essential status. For each response variable and interaction, we provide the t-value and the p-value. All response variables are scaled to zero mean and unit variance before model run.

| **Model** | **Variable** | **Distance traveled** | |
|---|---|---|---|
| | | ***t*** | ***p*** |
| **Mean value in phase iii** | Proportion poor | 11.1 | <0.001 |
| | Proportion essential | 7.59 | <0.001 |
| | Population density | -9.70 | <0.001 |
| | Proportion Black | -4.02 | <0.001 |
| | Proportion poor : proportion Black | 1.84 | 0.065 |
| | Proportion poor : proportion essential | -6.08 | <0.001 |
| | Proportion Black : proportion essential | 3.32 | 0.001 |

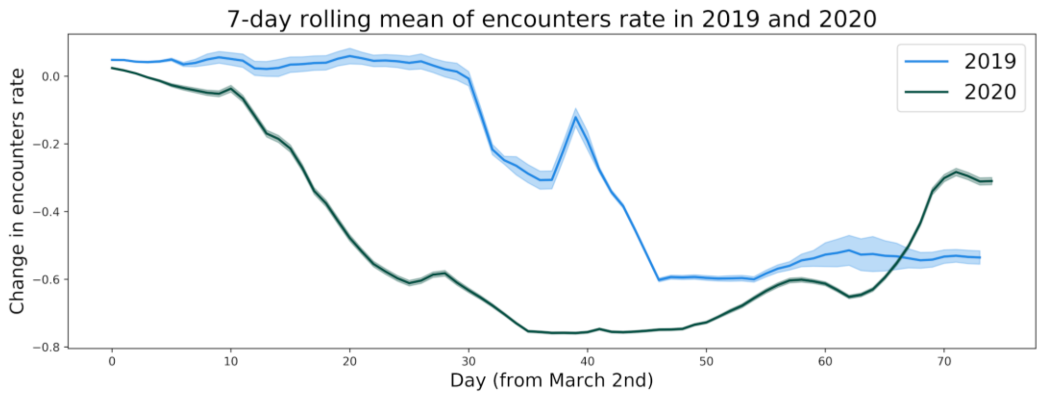


Figure S1: **The seasonality of encounters:** We show a 7-day rolling mean of the encounters rate from March 2nd onward for 2019 (blue) and 2020 (green). The solid line represents the mean and the shaded area two standard errors of the mean.
